# Automated estimation of frequency and spatial extent of periodic and rhythmic epileptiform activity from continuous electroencephalography data

**DOI:** 10.1101/2025.05.29.25328582

**Authors:** Alexandra-Maria Tăuţan, Jin Jing, Lara Basovic, Peter N. Hadar, Shadi Sartipi, Marta P. Fernandes, Jennifer Kim, Aaron F. Struck, M. Brandon Westover, Sahar F. Zafar

## Abstract

**Background and purpose:** Rhythmic and periodic patterns (RPP) are harmful brain activity observed on continuous electroencephalography (cEEG) recordings of critically ill patients. The presence of RPPs at higher frequencies and on a larger scalp area (spatial extent) are associated with a higher probability of poor outcomes. This work describes automatic methods for detection of the frequency and spatial extent of specific RPPs: lateralized and generalized rhythmic delta activity (LRDA, GRDA) and lateralized and generalized periodic discharges (LPD, GPD).

**Methods:** The frequency and spatial extent of RPPs is estimated using signal processing techniques combined with rule-based logic. The validation of the algorithms was performed on a total of 1087 cEEG segments. The annotations of three expert neurophysiologists for event frequency and spatial extent were considered the gold standard for the evaluation of the algorithm output. The inter-rater reliability (IRR) is evaluated for the assessment of performance.

**Results:** The selected algorithms match or exceed the agreement of experts on the frequency and spatial extent of RPP segments. RDA1b-FFT (Fast Fourier Transform), the best algorithm for rhythmic delta activity, showed an expert-algorithm IRR ranging from a good to excellent intra-class correlation coefficient (ICC) of 66-96%, whereas the expert-expert IRR ranged from 60-92%. The best algorithm for periodic discharges, PD2a, showed an expert-algorithm IRR ranging from ICC of 13-80%, whereas the expert-expert IRR ranged from 13-86%.

**Conclusions:** The proposed algorithms for estimating frequency and spatial extent of rhythmic and periodic patterns match expert performance and are a viable tool for large-scale cEEG analysis.

**Highlights:**

- Rhythmic and periodic epileptiform activity are harmful electroencephalographic (EEG) patterns observed in critically ill patients and are associated with poorer outcomes particularly at higher frequencies and spatial extent
- Automatic estimation of frequency and spatial extent would allow large scale EEG studies linking physiological patterns to treatments and outcomes
- We developed algorithms for the automatic quantification of frequency and spatial extent of epileptiform activity that match the performance of human annotators

## 1. Introduction

The last two decades have seen a significant increase in continuous electroencephalography (cEEG) utilization in the critical care setting [1]. cEEG is used for detection of seizures and other seizure-like rhythmic and periodic epileptiform patterns (RPPs) in at risk critically-ill and comatose patients [2], [3]. In addition to seizure detection, cEEG is also used for prognostication in patients with severe brain injuries and for detection of cerebral ischemia [4], [5], [6].

Up to half of critically-ill patients undergoing cEEG monitoring are found to have seizure-like RPPs that do not fall under the definition of seizures [7]. The American Clinical Neurophysiology Society (ACNS) proposed terminology to harmonize the interpretation of rhythmic and periodic patterns both for clinical care and research [8]. Periodic discharges are defined as repetitive waveforms with clear inter-discharge background activity occurring at regular intervals. Rhythmic delta activity is characterized by repetitive slower waves between 0.5-4Hz. ACNS further characterizes these patterns as lateralized or generalized periodic discharges (LPDs, GPDs) and lateralized or generalized rhythmic delta activity (LRDA, GRDA), and the ictal-interictal continuum (IIC). The IIC is defined as a combination of LPDs, GPDs or LRDA observed in 10 seconds of EEG recordings at specific frequencies and that do not qualify as seizures. Fig. 1 shows examples of lateralized and generalized periodic and rhythmic activity.

**Figure 1.**
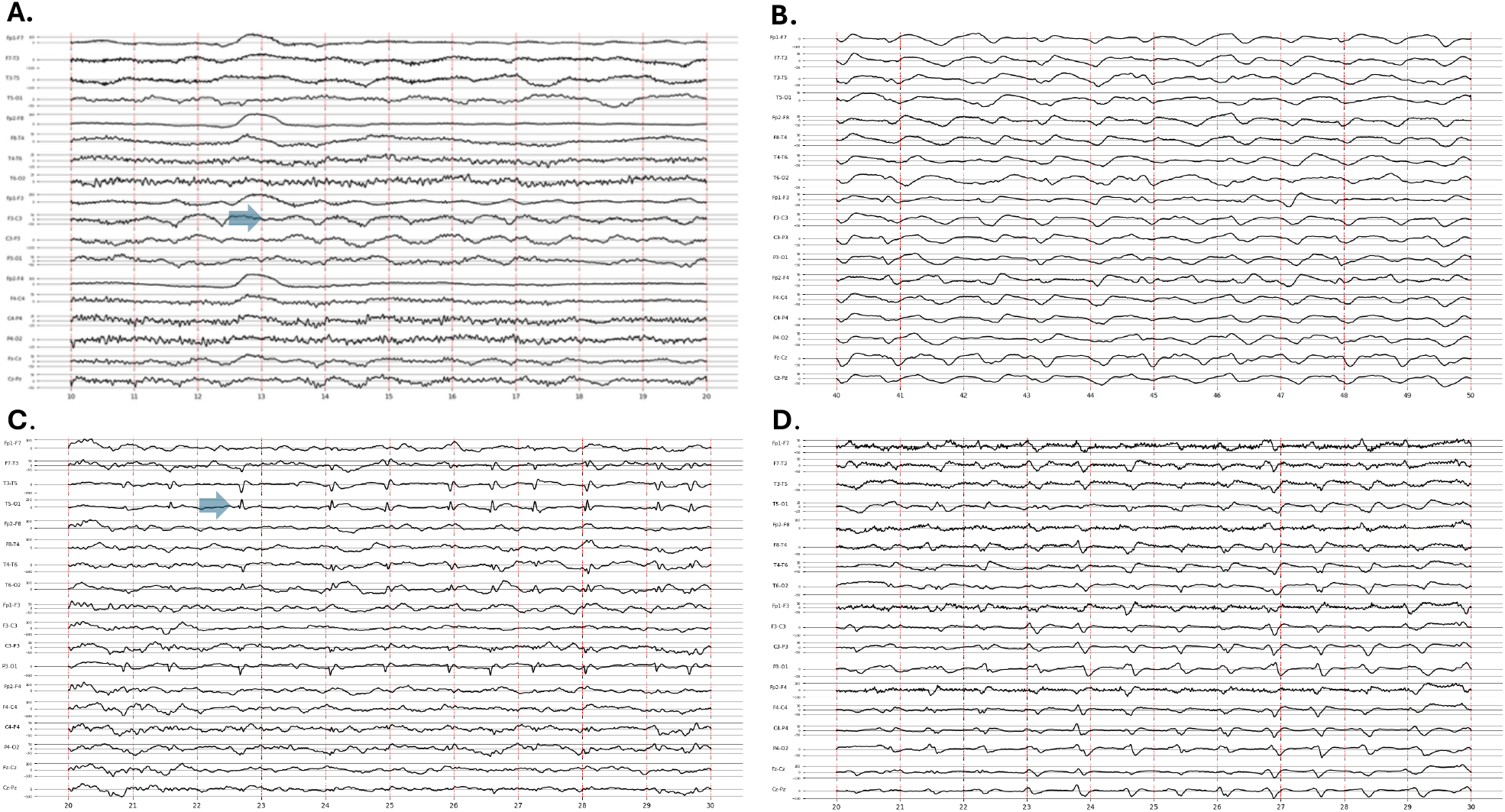
Examples of Periodic and rhythmic patterns. A. Lateralized rhythmic delta activity (LRDA) with a blue arrow highlighting an example of an event. B. Generalized rhythmic delta activity (GRDA). C. Lateralized periodic discharges (LPD) with a blue arrow highlighting and example of an event. D. Generalized periodic discharges (GPD)

RPPs and the IIC have been directly linked to higher likelihood of mortality and worse functional and cognitive outcomes in patients with acute brain injuries such as stroke and trauma [9], [10], [11]. In particular, higher frequency and spatial extent of these patterns are linked to worse outcomes [3]. The frequency of RPPs and spatial extent of brain regions involved have been associated with a monotonic increase in metabolic activity as demonstrated by metabolic and functional imaging and cerebral microdialysis analysis [12]. Several studies have shown that presence of LPDs, particularly at frequencies higher than 2Hz, is linked to an increased risk of seizures [13], [14] and increased mortality [15]. GPDs have shown an equivalent seizure risk to LPDs when present at frequencies higher than 1.5Hz [2]. LRDA has a similar frequency-dependent relation with seizures and worse outcomes [16], while GRDAs are the least harmful of the patterns with no association to seizure risks [13]. Hence both the frequency and spread of the pattern (lateralized or generalized) may be helpful in identifying patients at risk of a poorer outcome. Furthermore, precise localization of these patterns in particular lobes can indicate the underlying cause of the abnormal pattern. For instance, temporal RDAs (TRDA) can be an indication of focal epilepsy or of mesial temporal atrophy [17].

While RPPs and the IIC have been linked to worse outcomes, there is limited data to guide anti-seizure treatment approaches [2]. One of the major limitations in pursuing large-scale clinical research studies to examine the impact of anti-seizure treatment is the resource and labor-intensive task of manual review of hours of EEG signals collected in critical care settings. Furthermore, there are still controversies related to best treatment approaches, and their link to IIC patterns and outcomes [2]. Automatic methods of identifying RPPs and IIC patterns and estimating their frequency and spatial extent, can help with clinical patient monitoring as well as conducting research studies on large patient populations to identify links between treatments and outcomes.

Prior work on the automatic annotation of EEG signals has focused primarily on classification of RPPs and IIC patterns [18], [19], [20], [21]. Several other works have examined the presence or frequency of RPPs but have been limited by sample sizes or restricted only to one pattern type and have not quantified spatial extent [18], [20], [22], [23], [24]. This work aims to advance state-of-the-art algorithms for frequency and spatial extent estimation of RPPs and IIC patterns by (i) proposing algorithms to calculate the frequency and spatial extent of LRDA, GRDA, LPD and GPD EEG segments, (ii) comparing agreement between the algorithm and expert EEG readers with the inter-rater agreement between experts in estimating the frequency and spatial extent of epileptiform patterns, (iii) proposing a validation framework for frequency and spatial extent estimations.

## 2. Methods

### 2.1. Dataset and annotation protocol

This study was conducted under protocols approved by the institutional review boards of Massachusetts General Hospital (protocols #2023P000487, #2024P002630) and Beth Israel Deaconess Medical Center (protocols #2022P000481, #2022P000417), which waived the requirement for informed consent for this retrospective analysis. We used a dataset comprised of 10 second EEG recordings previously labelled as LPDs, GPDs, LRDAs, and GRDAs by 30 clinical experts trained in neurophysiology as described in Jing et al [25]. From the dataset, EEG segments were selected for further analysis of frequency and spatial extent based on degree of consensus on pattern classification. We selected segments where majority of annotators agreed on a single pattern type. A total of 296 LPD, 296 GPD, 210 LRDA and 285 GRDA segments were selected for further annotation of frequency and spatial extent. Each segment contained 19 raw EEG channels sampled at 200Hz. Annotations were conducted on 18 differential channels re-referenced to a longitudinal bipolar montage. Three clinical neurophysiologists (SFZ, PH, LB) performed independent annotations. Given the inter-rater reliability for classification in the original dataset was a moderate percent agreement of 52% [25], the three annotators initially reviewed the segments for pattern classification. Only those segments with 100% agreement on the classification between all three annotators were used for frequency and spatial extent analysis. A final set of 50 LRDA, 119 GRDA, 111 LPD and 109 GPD segments were selected for the performance evaluation of the frequency and spatial extent algorithms. As the developed algorithms are rule-based, no training set was needed.

The labeling of the selected 10 second EEG segments was performed using a custom interface that allowed annotators to enter the frequency of the event and spatial extent (see Fig. A1 from the supplemental material). The reported frequency of event was the mean value observed within the 10 second segment. Spatial extent was reported as the number of channels that have events (0 – no channels showed an event, 1 – all 18 channels showed the event).

### 2.2. Frequency and Spatial Extent Detection

An overview of the event detection algorithm is provided in Fig 2. The initial 10s segment of the EEG recording was re-referenced to a longitudinal bipolar montage. A total of 18 differential channels are available. Preprocessing was performed by applying a 60Hz notch filter and a 0.5-40Hz bandpass filter [26]. Different algorithms were used for RDA and PD events as the information in these phenomena is encoded differently in the recorded EEG signals. RDAs are predominantly characterized by the frequency content of the repetitive waveforms, while PDs are characterized by morphology of the discharges in addition to their repetitive nature. Thus, we base methods for detecting frequency and spatial extent for RDAs on frequency domain transformations, whereas for PDs we base our algorithms on time domain signal processing. For spatial extent, each channel was analyzed individually. Several methods for RDA and PD detection are proposed and evaluated:

- RDA 1a-b – Fourier Transform based
- RDA 2 – Hilbert Huang Transform based
- PD 1 – Prior Detector - McGraw et al [22]
- PD 2a-b – Derivative Peak Detection based

**Figure 2.**
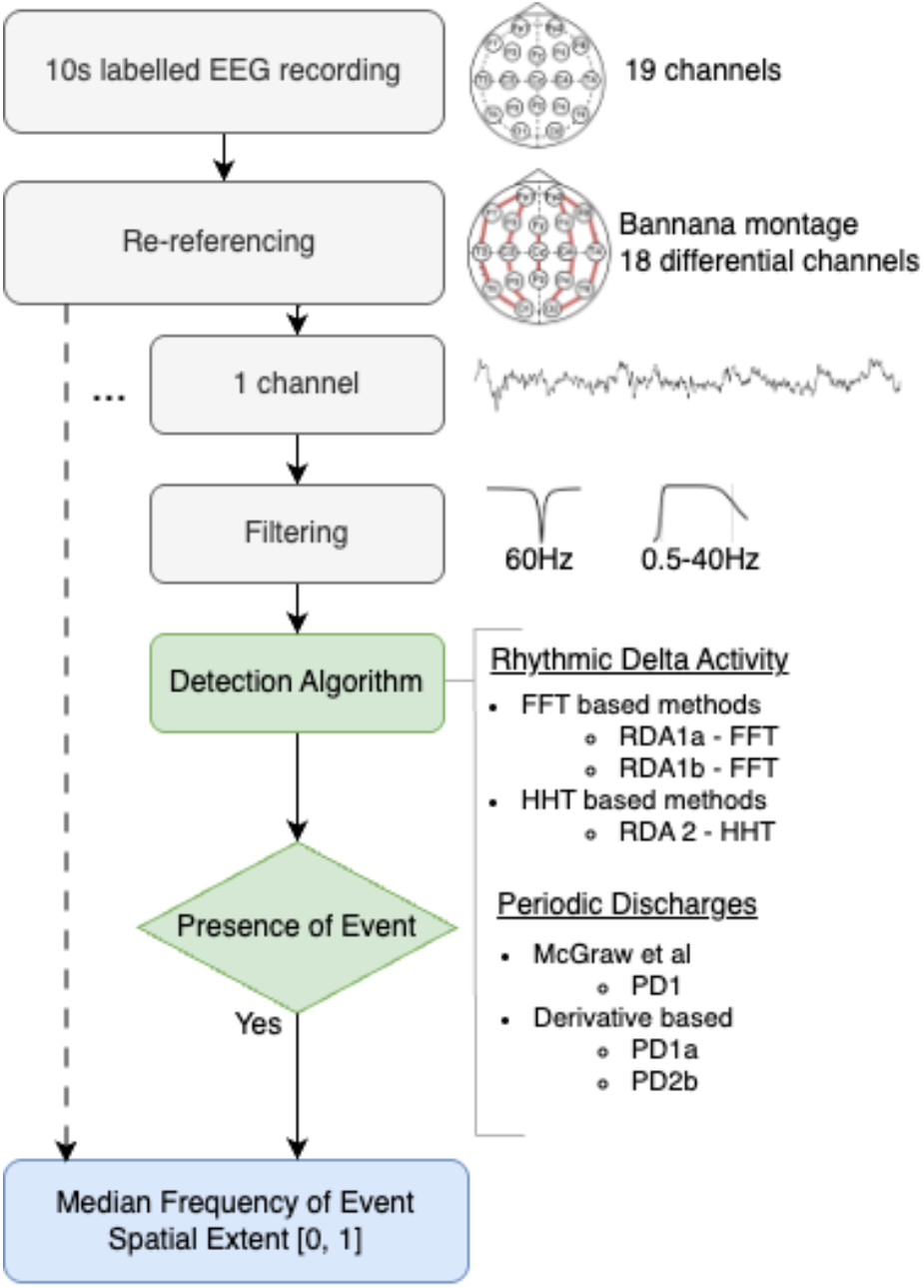
Overview of the methods applied for detecting frequency and spatial extent of epileptiform activity. Detection of PDs and RDAs are approached diKerently through several algorithms. PD – periodic discharges, RDA – rhythmic delta activity, FFT – Fast Fourier Transform, HHT – Hilbert Huang Transform.

These algorithms are detailed in the following sections. The presence or absence of the event along with the corresponding frequency were obtained for each of the differential channels. The output of the algorithm was the median frequency of event across all channels, the spatial extent as a ratio between the number of channels with an event and the total number of channels, and the spatial areas defined based on the channels that show an event.

#### 2.2.1. RDA Detection

Three frequency-based methods were explored for detection of rhythmic delta activity. Schematics of the developed algorithms are illustrated in Fig. 3. The first two, RDA 1a and RDA 1b, are variations based on the Fourier Transform of the EEG, while the third RDA 2 is based on the Hilbert-Huang Transform.

**Figure 3.**
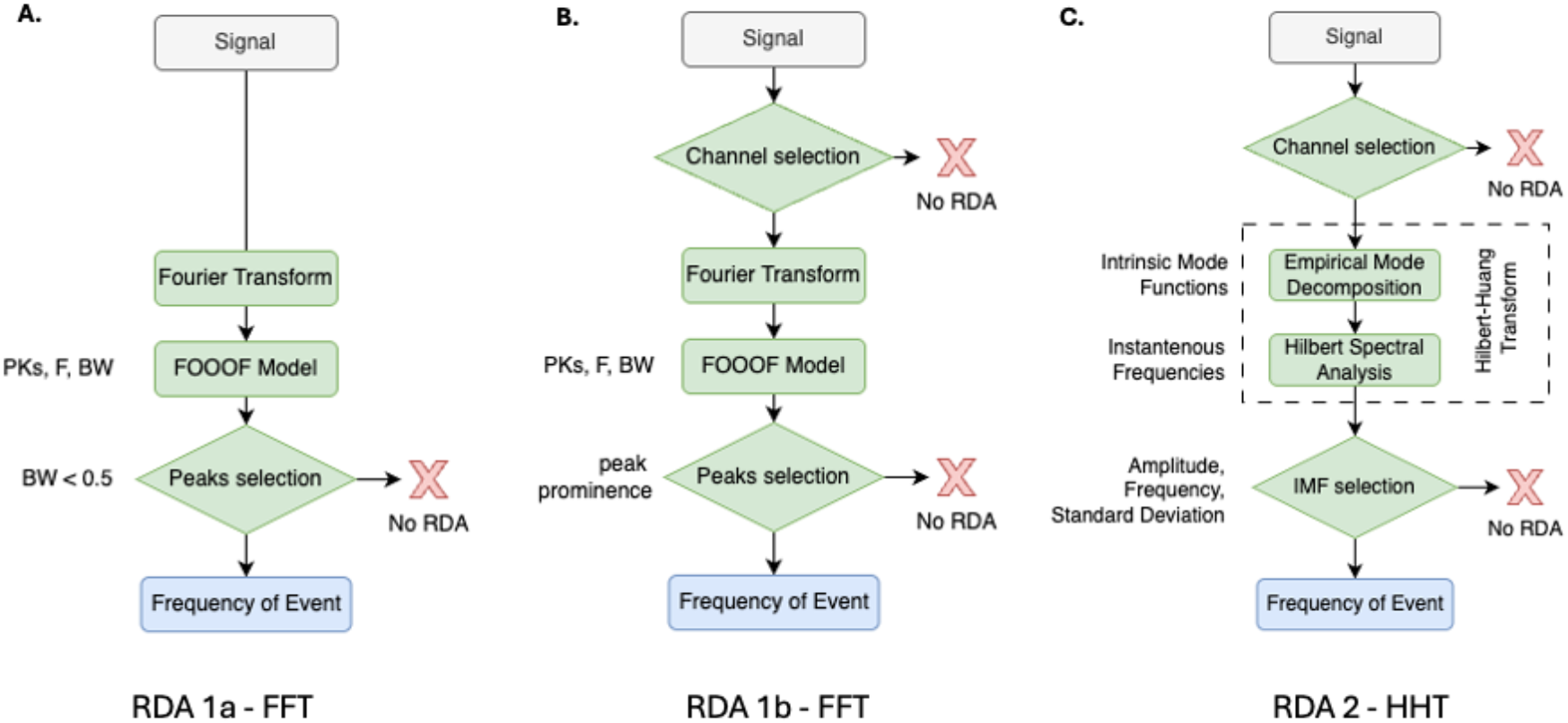
Schematics of the three algorithm variations for the detection of rhythmic delta activity frequency and spatial extent. A.B. Detection methods based on the Fourier Transform; C. Detection method based on the Hilbert Huang Transform. RDA - Rhythmic delta activity, FFT – Fast Fourier Transform, HHT – Hilbert-Huang Transform, FOOOF – Fitting oscillations and one over f, PKs – adjusted power of peaks, F – center frequency, BW – peak bandwidth, IMF – Intrinsic Mode Function.

##### 2.2.1.1. RDA1 - Fourier Transform Based

The power spectral density (PSD) of the 10s EEG segment was calculated based on the Fourier Transform:

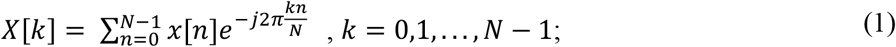

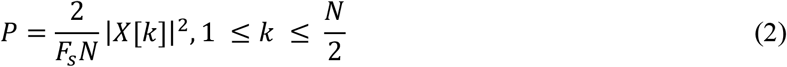

where P is the power spectral density, *F*_*S*_ is the sampling frequency, *N* the signal length, *X*[*k*] the Fourier transform of the discrete *x*[*n*], *x*[*n*] the input EEG signal, *n* the number of samples, *k* the frequency components.

The PSD of the EEG is characterized by both periodic and aperiodic components [27]. In case of RDAs, the aim is to detect periodic components represented as narrow band peaks in the 0.5-4Hz range. The power spectrum of EEG data at low frequencies is characterized by a high power aperiodic (1/f like) component that overlaps with the target delta peaks and can compromise their detection. To address this challenge, a model fitting the aperiodic component as a Lorentzian function and the periodic components with Gaussians (*fooof*) as described by Donoghue et al [28] is applied:

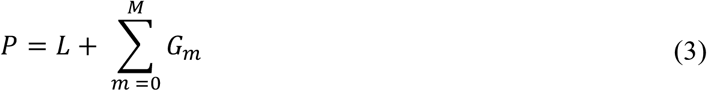

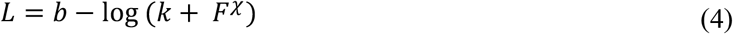

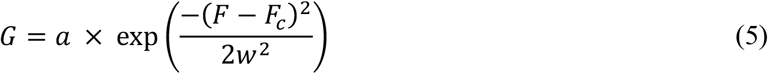

where L is the aperiodic component, G represents the Gaussian function and M the total number of Gaussians, *b* the broadband offset, *χ* the exponent, *k* the parameter controlling the bend of the aperiodic component, *F* the vector of frequencies, *a* is the peak power, *F*_*c*_ is the center frequency and *w* is the standard deviation of the Gaussian.

The output of the model provides the central frequency (*F*_*c*_), the aperiodic-adjusted power (PK) and the bandwidth (*BW*) of detected peaks.

###### RDA1a-FFT

The first RDA detection algorithm is described in Fig.2a. It takes the output of the *fooof* model and selects narrow band peaks by considering the condition *BW* ≤ 0.5.. If no peaks are detected or selected by applying the narrow-band condition, no RDA event is detected. If one or more peaks meet the condition, the frequency of event is considered as the mean central frequency of the detected peaks.

###### RDA1b-FFT

The second RDA detection algorithm is a variation of the first and is described in Fig.2b. Prior to applying the Fourier Transform, a channel selection is applied by identifying channels with invalid EEG data. Channels with corrupt EEG data are common in clinical recordings and can be caused by a variety of reasons such as motion artifacts, increased skin-electrode contact impedance or wire disconnections. The following condition is applied to identify channels with valid data:

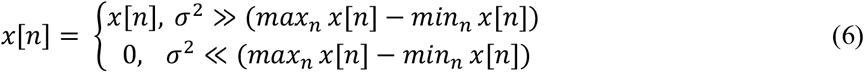

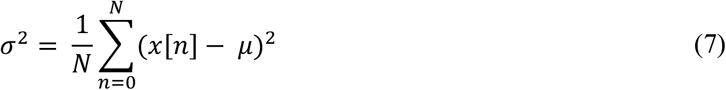

where *σ*^2^ is the variance of the signal, *max*_*n*_ *x*[*n*] - *min*_*n*_ *x*[*n*] is the range of the signal, N is the signal length and *μ* is the mean.

If the variance of the signal was not significant compared to the signal range, we conclude that the channel does not contain a valid EEG signal. If the channel did not contain valid EEG data, we conclude that no RDA was present. Channels with valid EEG data were further processed using *fooof* modelling as described previously. When selecting narrow band peaks representatives of RDA, selection was enhanced by considering the prominence of the peak with respect to the mean spectrum in the band of interest as an additional control. For each detected peak, *P*_*k*_, the band of interest was defined as:

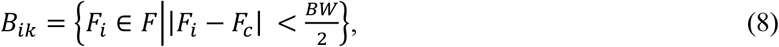

while the baseline frequency band is considered:

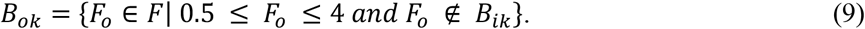

A peak is selected when:

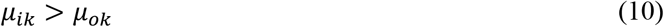

where *μ*_*ik*_ and *μ*_4*k*_ is the mean power spectral density for the peak band of interest and baseline. If one or more peaks meet the conditions applied, the frequency of event was considered as the mean central frequency *F*_*c*_ of the selected peaks.

##### 2.2.1.2. RDA2 - Hilbert-Huang Transform Based

The Hilbert-Huang Transform (HHT) is an effective tool for analyzing the time-frequency content of non-stationary and nonlinear data [29]. It has been successfully applied in the analysis of EEG data for a variety of tasks including movement identification [30], evaluating connectivity metrics [31] and mental imagery tasks [32]. The HHT is composed of two parts: empirical mode decomposition (EMD) and Hilbert spectral analysis (HSA). By applying EMD to the original signal *x*[*n*], a set of sub-signals *c*_*i*_[*n*] called Intrinsic Mode Functions (IMF) are obtained. The IMFs represent an adaptive orthogonal basis for the signal which can be described as:

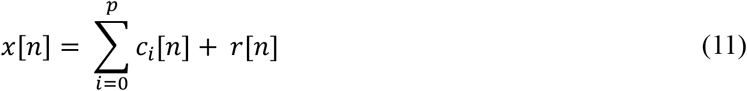

where *r*[*n*] is a residual component representing a trend or a constant. All IMFs satisfy two conditions: (i) the number of extrema and zero-crossings can differ by maximum one, (ii) the mean value of the upper and lower envelopes must be zero. The Hilbert Transform (HT) is applied to all IMFs to obtain their instantaneous frequencies [33]. The HT of the IMFs is calculated as follows:

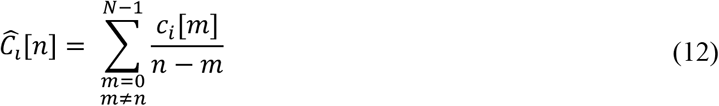

With the HT of the IMFs, the instantaneous amplitude and frequency can be obtained from the analytical signal by combining 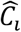 [*n*] and the original signal:

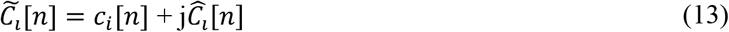

The module of 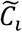 is the instantaneous amplitude:

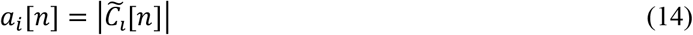

The instantaneous frequency can be obtained by deriving the phase of the analytical signal:

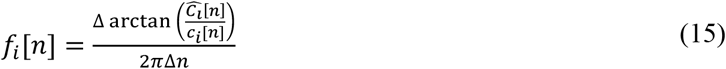

The Hilbert Spectrum is characterized by both the instantaneous amplitude and frequency functions [*a*_*i*_[*n*], *f*_*i*_[*n*]].

###### RDA2-HHT

The third RDA detection algorithm is described in Fig. 2c. Valid EEG data was identified on the channel by applying the conditions identified in Eq. (6). If there was corrupt EEG data, the channel was marked as having no RDA. If the data was valid, the HHT was applied by calculating the IMFs through empirical mode decomposition and obtaining the instantaneous frequencies through the Hilbert Transform. Not all information contained in the HSA is relevant to the detection of RDA components. Previous methods focused the selection of IMFs based on statistical properties, periodicity degree or information shared with the original signal [34], [35]. A series of conditions were applied to select relevant IMFs based on the statistical properties of their amplitude and instantaneous frequency content, as well as the proportion of the IMF amplitude with respect to the original signal:

- 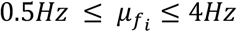, where 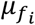 is the mean of the instantaneous frequency of *c*_*i*_
- 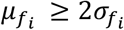, where 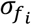 is the standard deviation of the instantaneous frequency of *c*_*i*_
- 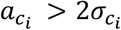, where 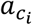 is the maximum amplitude of *c*_*i*_, and 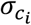 its standard deviation
- 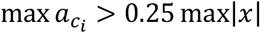

If all the conditions were valid, the IMF is selected. The frequency of the RDA event was considered the mean of the instantaneous frequencies of all the selected IMFs.

#### 2.2.2. PD Detection

The detection of periodic discharges was approached through the identification of events in the time domain. We made use of the algorithm presented by McGraw et al [22] and integrated it into our event detection framework. Additionally, we proposed a new detection method by shifting the event detection to the first derivative of the EEG signal and applying two different peak detectors.

##### 2.2.2.1. PD 1 - Prior PD detector

McGraw et al [22] proposed an algorithm for LPD and GPD frequency of event and spatial extent detection. After pre-processing, a high-low percentile thresholding is applied on the time domain EEG signal to select peaks that are potential PDs. Fixed thresholds on the amplitude and absolute values are applied on the identified peaks to filter out false detections. Once peaks are identified for the entire recording, the spatial extent is calculated based on the global or local energy of the detected events and reported as the ratio with respect to the total number of channels. The frequency of the event is reported as the median inter-discharge interval frequency at the channel group level.

##### 2.2.2.2. PD 2 - Derivative Peak Detection Based

With the goal of reducing false detection rates, we developed an alternate PD detector with the initial event identification based on the first derivative of the EEG signal. An overview of the proposed detection method is available in Fig. 4.

**Figure 4.**
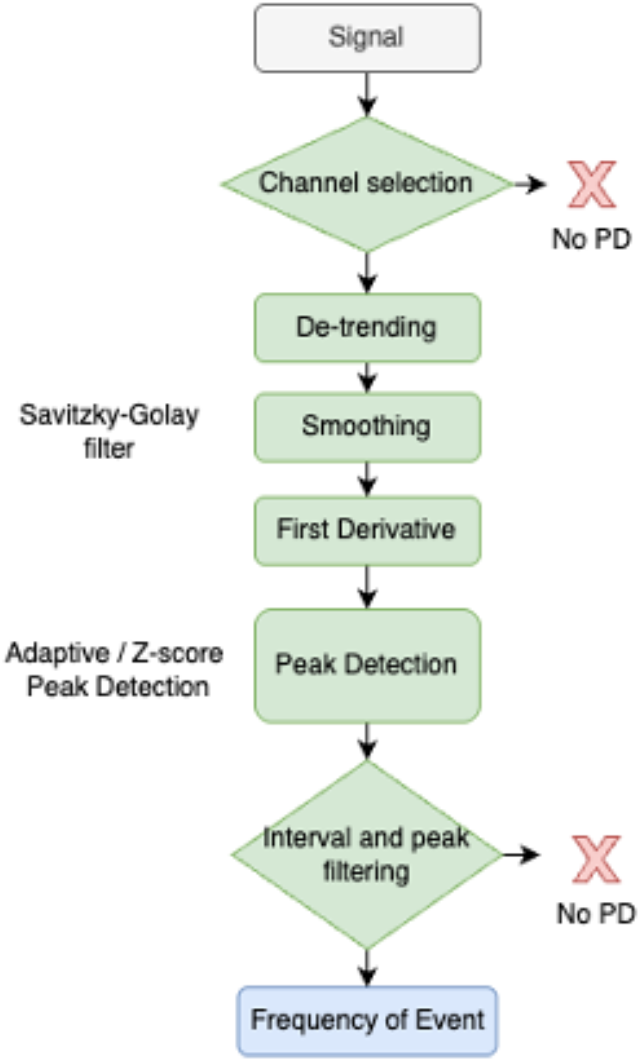
Schematic of the derivative based PD (periodic discharges) detector algorithms. Two alternative algorithms are used for peak detection based on the signal derivative.

Periodic discharges are often characterized by abrupt changes in the amplitude of the EEG signal. The derivative of a signal captures the rate of change in the original signal. Slopes in the original signal correspond to peaks of the first derivative. A higher rate of change in the original signal is reflected as a high amplitude peak in the first derivative. Thus, the detection of periodic discharges can be performed by detecting corresponding peaks in the first derivative of the EEG signal.

The detection of peaks is essential in processing a variety of physiological recordings [36], however peak detection is challenging due to background noise and interference. To eliminate potential sources of interference and isolate the PD events from other EEG phenomena, several pre-processing steps were applied. First, the channel was evaluated for containing valid EEG data as described previously in Eq. (6). Next, the signal was detrended by subtracting the mean and smoothed by applying a Savitsky-Golay 2^nd^ order polynomial filter on a 10-sample window [37]. Afterwards the first derivative of the EEG signal was computed. Two methods for peak detection were evaluated as described below.

**PD2a**. The first makes use of local adaptive thresholds based on the statistics of the first derivative signal. A peak was identified when a local maximum is above a locally defined statistical based threshold within a 1 second sliding window as defined below:

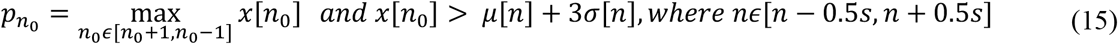

**PD2b**. The second peak detector explores the use of a z-score signal with peaks identified on a locally derived threshold:

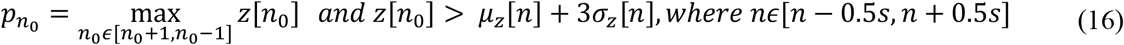

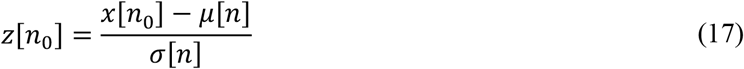

where *z* is the z-score of the signal in the defined 1 second window, *μ*_F_ is the mean of the z-score signal, *σ*_F_ is the standard deviation of the z-score signal.

After applying either of the peak detectors, the peaks identified were further filtered based on the length of the intervals between the peaks.

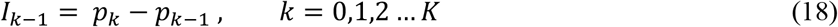

where *I* is the inter-peak interval, *p*_*k*_ is the timing of peak *k* and *K* is the total number of peaks detected. A channel is considered to have PD events, if:

- More than one inter-peak interval is identified: *K* > 2
- The detected inter-peak intervals are uniformly spaced with *σ*_G_ < *i*, where *σ*_G_ is the standard deviation of the *I* vector

The inter-peak intervals were divided by the sampling frequency and the average was reported as the frequency of the event.

### 2.3. Evaluation method

The gold standard for assessing frequency and spatial extent of LRDA, GRDA, LPD and GPD epileptiform activity were the manual annotations of EEG recordings by experienced clinicians. We evaluated the output of the proposed detection methods by comparing to manual annotations of the EEG segments by experts.

We examined the inter-rater reliability between experts (ee-IRR) with the agreement between the algorithms and experts (ea-IRR). The annotations for the frequency and spatial extent of events as well as the output of the algorithms were evaluated through the intra-class correlation coefficient (ICC) [38].

Additionally, the continuous frequency and spatial extent data was categorized to allow reporting of mean percentage agreement (PA). The categorization was performed as follows:

- Frequency of event intervals: <1 Hz, 1-1.5Hz, 1.5-2Hz, 2-2.5Hz, 2.5-3Hz, >3Hz
- Spatial extent intervals: 1-4 channels, 5-10 channels, 11-14 channels, 15-18 channels

Inter-rater agreement for both ICC and PA were quantified as excellent if the values are 75% - *ikk*%, good for 6*k*% - 74%, fair for 4*k*% - 59% and poor for values < 4*k*% [39].

The output of the algorithms was further evaluated by calculating the mean absolute error (MAE) between the annotated values and the calculated ones:

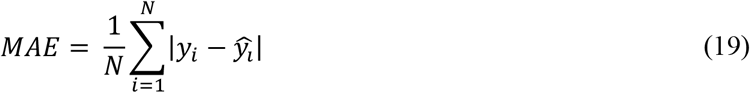

where N is the total number of segments, *y*_*i*_ is the annotated values, 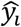 is the output of the algorithms.

## 3. Results

The results for the ICC and mean PA between the annotators and the proposed algorithms are available in Fig.5 for RDA and Fig. 6 for PD.

**Figure 5.**
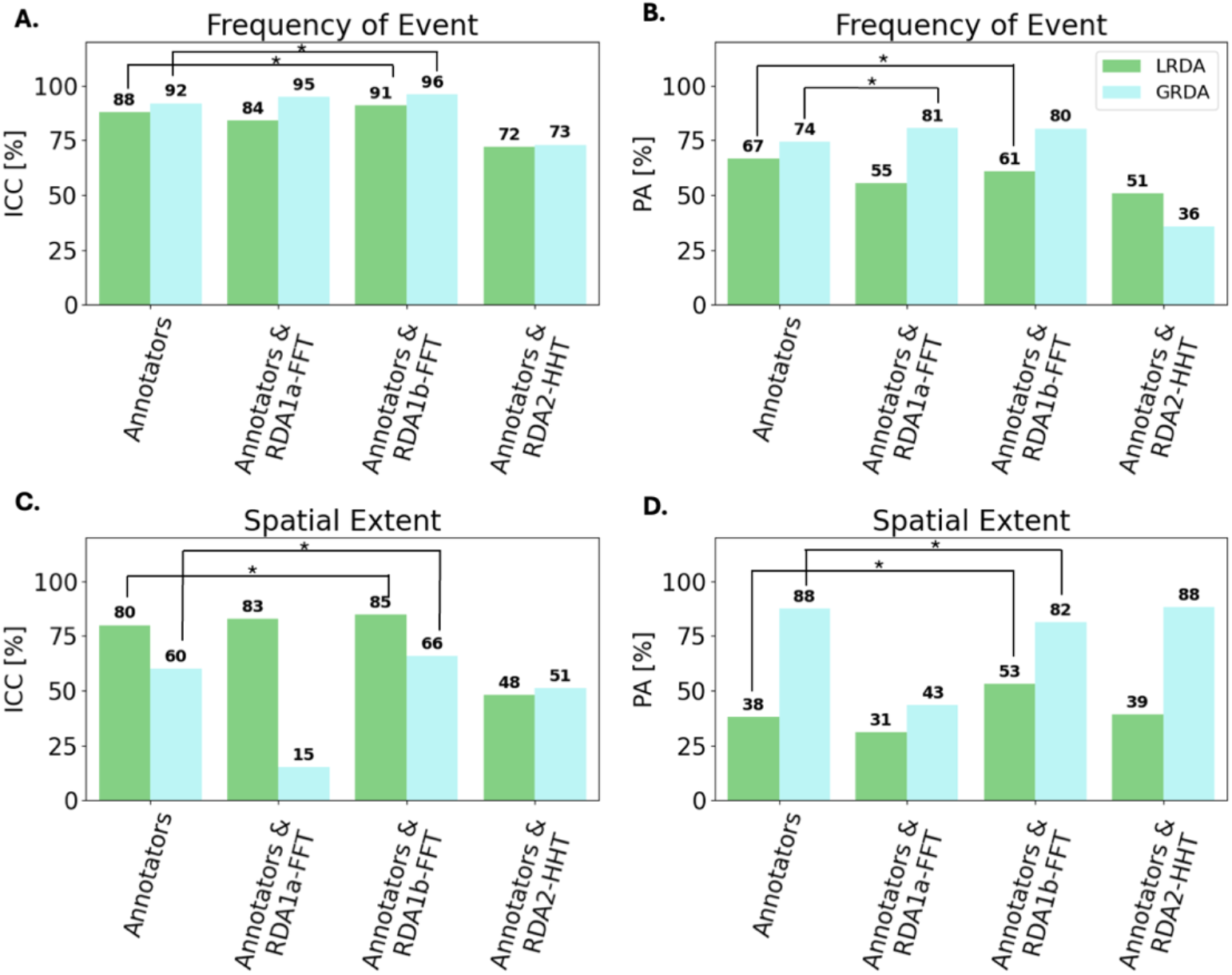
Intra-class correlation coeKicient (ICC) and mean pair-wise agreement (PA) between annotators and algorithms for LRDA and GRDA segments. The star indicates the best performing algorithm with an ea-IRR (expert-algorithm inter-rater reliability) that is higher when compared to ee-IRR (expert-expert inter-rater reliability). A. ICC for frequency of event. B. PA for frequency of event. C. ICC for spatial extent. D. PA for spatial extent.

**Figure 6.**
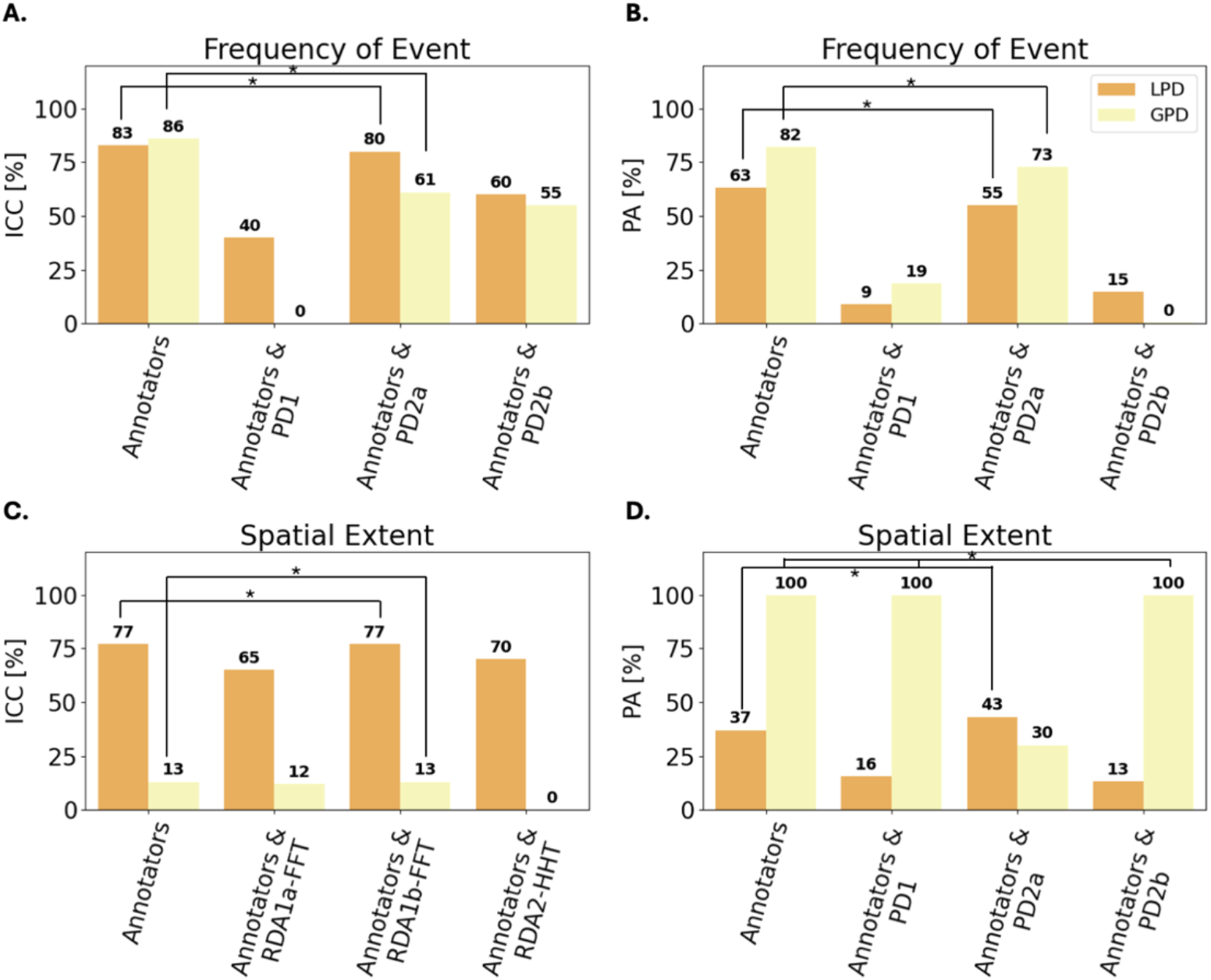
Intra-class correlation coeKicient (ICC) and mean pair-wise agreement (PA) between annotators and algorithms for LPD and GPD segments. The star mark indicates the best performing algorithm with an ea-IRR (expert-algorithm inter-rater reliability) that is comparable or better than ee-IRR (expert-expert inter-rater reliability). A. ICC for frequency of event.B.PA for frequency of event. C. ICC for spatial extent. D. PA for spatial extent.

The IRR analysis shows that the output of the RDA1b-FFT algorithm is comparable to that of human annotators. The ea-IRR [95% confidence interval] ICC for RDA1b-FFT, was 91% [CI 85,94] and 96% [CI 94,97] for LRDA and GRDA respectively. The mean pair-wise agreement for LRDA was 60.66% [47,72]. RDA1b-FFT scored lower for GRDA with a difference of 0.3% from RDA1a-FFT. The highest algorithm ea-IRR for spatial extent was also obtained by RDA1b-FFT with an ICC 85% [CI 77,91] and 66% [CI 46,71] for LRDA and GRDA respectively, and a mean pair-wise agreement of 53% [CI 39,64] for LRDA. Spatial extent for GRDA was best for RDA2-HHT, ~2% higher than RDA1b-FFT. A detailed overview of the IRR analysis is available in Tables B1-2 from the supplementary materials.

For calculating the frequency and spatial extent of PDs, the PD2a algorithm showed the highest performance within the same range as the performance of human annotators. The ea-IRR ICC for frequency of LPD and GPD segments, for algorithm PD2a was 80% [CI 73,85] and 61% [CI 47,72] and a mean pair-wise agreement of 54.95% [CI 45,63] and 72.78% [CI 74,88] for LPD and GPD respectively. The calculated spatial extent also showed the highest ea-IRR for PD2a with an ICC of 77 % [CI 70,84] and 13% [CI 17,37] for LPDs and GPDs respectively, with a percentage agreement of 43.20% [CI 34,52].

In supplemental Fig.C1-4 we show the relation of raw pair-wise agreement of the annotators and algorithms, as well as scatter plots of the annotated values for frequency and spatial extent. While the plots show a significant variation in the comparison of the algorithms’ output and the various annotators, the reported R^2^ is consistently higher for RDA1b-FFT with respect to all annotators and in close range to the annotator-annotator R^2^ when compared to the values for RDA1a-FFT and RDA2-HHT. Similarly, PD2a shows a consistently higher R^2^ which aligns with the performance reported via ICC and PA.

The mean absolute error between the mean value reported by the annotators and the outputs of each algorithm are shown in Table 1. The mean absolute error for the entire dataset, including segments with rater disagreement on classification is available in Table B3 of the supplemental materials. RDA1b-FFT and RDA2-FFT had the lowest mean absolute error (0.13Hz) for LRDA frequency. RDA1a-FFT had the lowest mean absolute error (0.24Hz) for GRDA frequency. RDA2-HHT had the lowest mean absolute error (0.14) for LRDA spatial extent and RDA1b-FFT had lowest error (0.09) for GRDA spatial extent. PD2a had the lowest mean absolute error for LPD (0.41Hz) and GPD (0.15Hz) frequency, as well as the lowest error for LPD spatial extent (0.17). PD2b had the lowest mean absolute error for GPD (0.01) spatial extent.

**Table 1.**
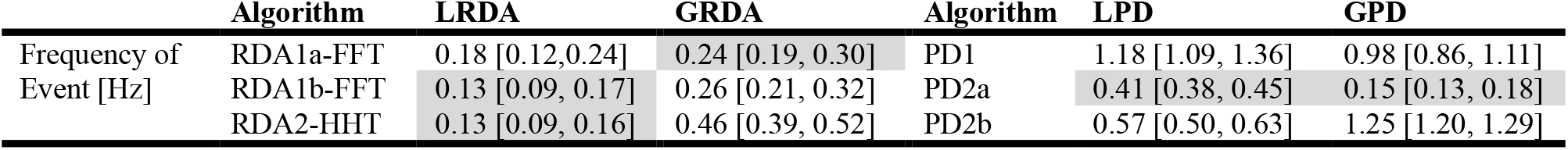
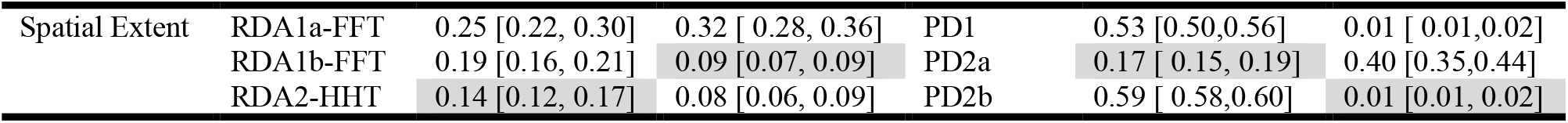
MAE (mean absolute error) between the mean human expert annotated value and the algorithms for RDA (rhythmic delta activity) and PD (periodic discharges), along with their 95% CI. The lowest values for each event type are highlighted.

|  | Algorithm | LRDA | GRDA | Algorithm | LPD | GPD |
| --- | --- | --- | --- | --- | --- | --- |
| Frequency of Event [Hz] | RDA1a-FFT | 0.18 [0.12,0.24] | 0.24 [0.19, 0.30] | PD1 | 1.18 [1.09, 1.36] | 0.98 [0.86, 1.11] |
|  | RDA1b-FFT | 0.13 [0.09, 0.17] | 0.26 [0.21, 0.32] | PD2a | 0.41 [0.38, 0.45] | 0.15 [0.13, 0.18] |
|  | RDA2-HHT | 0.13 [0.09, 0.16] | 0.46 [0.39, 0.52] | PD2b | 0.57 [0.50, 0.63] | 1.25 [1.20, 1.29] |
| Spatial Extent | RDA1a-FFT | 0.25 [0.22, 0.30] | 0.32 [0.28, 0.36] | PD1 | 0.53 [0.50,0.56] | 0.01 [0.01,0.02] |
|  | RDA1b-FFT | 0.19 [0.16, 0.21] | 0.09 [0.07, 0.09] | PD2a | 0.17 [0.15, 0.19] | 0.40 [0.35,0.44] |
|  | RDA2-HHT | 0.14 [0.12, 0.17] | 0.08 [0.06, 0.09] | PD2b | 0.59 [0.58,0.60] | 0.01 [0.01, 0.02] |

Examples of the output from the best performing algorithms RDA1b-FFT and PD2a are shown in Fig. 7. The left side displays the filtered EEG signals, with detected RDA channels marked in blue and detected PD events marked in red. The right side of the panel details the estimated frequency of event, spatial extent and spatial areas.

**Figure 7.**
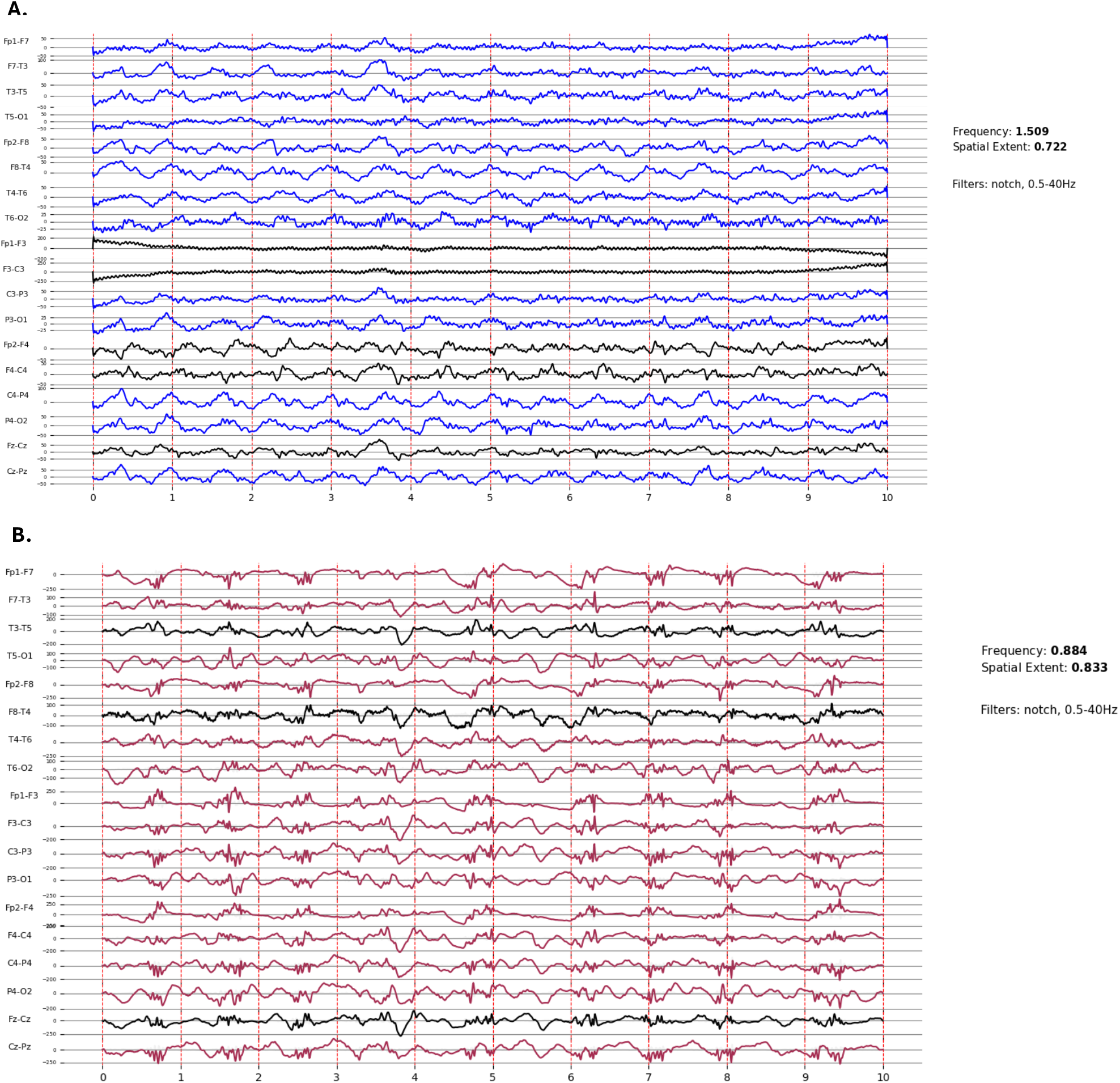
Output of the frequency of event and spatial extent. The EEG channels evaluated are displayed on the left, while the algorithm outputs are listed on the right. A. PD2a - red colored EEG traces represent channels with detected periodic discharges (PD). B. RDA1b-FFT – blue colored EEG traces represent channels detected by the algorithm as containing rhythmic delta activity (RDA).

## 4. Discussion

In this work we developed algorithms to estimate the frequency and spatial extent of epileptiform activity that perform comparably to human expert annotators. These algorithms can be applied to large neurophysiologic datasets for automated annotation of RPP frequency and spatial extent, after the classification into IIC patterns by specialized models [24].

The FFT-based methods designed for RDA segments scored similarly in terms of detection of event frequency, with little variation in corresponding ICC and PA. This is expected, as similar underlying frequency extraction methods are applied. Notably, RDA1a-FFT had lower performance in terms of spatial extent as compared to RDA1b-FFT that had an additional valid channel identifier. This underscores the importance of eliminating problematic EEG data and appropriately pre-processing the signals prior to applying detection algorithms. Across the frequency of event and spatial extent for rhythmic delta activity, the best performance was obtained by RDA1b-FFT. The RDA2-HHT had the lowest performance across the three algorithms. While the Hilbert-Huang Transform is a powerful method for representing frequency content, the rule-based selection of IMF and HSA components may introduce variability in the output across different data, potentially leading to reduced performance.

In case of PD detection, the frequency estimation is dependent on correct identification of peaks in single EEG channels. PD2a achieved the highest performance in terms of ICC, PA and MAE. Applying peak detection to the first derivative of the EEG signal, rather than to the raw signal, led to improved event detection rates. PD2a utilized a locally computed threshold for peak identification, resulting in better performance compared to PD2b, which relied on a global threshold applied across the entire signal. Threshold-dependent detectors are highly sensitive to the empirically chosen threshold values. Depending on the characteristics of the data, this can lead to either over-detection of signal fluctuations or omission of relevant events.

The MAE relative to the average human-annotated values ranged between 0.13-0.41Hz for the frequency of events detected by the best performing algorithms. These values fall within the estimation range reported by human-annotators, who typically use a maximum granularity of 0.25Hz to report event frequency.

Overall, both the MAE and the ICC had greater variability for spatial extent with lower agreement both for the human annotators and for human-algorithm comparison. The MAE for the best performing algorithm ranged between 0.14-0.40 which translates to 3 to 7 channels out of 18 with missed detection. Spatial extent has shown an overall lower IRR for human annotators as well. The highest MAE of 0.40 for the spatial extent identified by PD2a was reported for the GPD segments. Although the algorithm performs poorly in detecting spatial extent for GPD segments, the spatial extent detection can be corrected by factoring in prior classification into generalized activity [24]. Handling of imbalanced data with skewed distributions is a well-known limitation of inter-rater reliability metrics such as the intra-class correlation coefficient [40], [41]. The distribution of spatial extent annotation for generalized activity is skewed towards 1 as most EEG channels are expected to be involved. This leads to an unexpectedly low ICC, while the PA shows an almost perfect agreement (see Fig. 5 and 6).

The current state-of-the-art EEG annotating algorithms focus primarily on classification of RPPs and IIC patterns [18], [19], [20], [21]. Jing et al. [21] developed SPaRCNet - a deep neural network trained on more than 50000 ten second EEG segments labeled by multiple neurophysiologists to identify seizures, LPDs, GPDs, LRDAs, GRDAs and data that falls into neither of these classes. The performance of the model is in the same range as expert-level inter-rater agreement; however, it only indicates the classification of EEG segments into RPP and not their frequency or spatial extent. In this work, we have further expanded the characterization of these patterns in terms of frequency of events and spatial extent. Fürbass et al. [18] proposed a rule-based peak detection algorithm for identification of RDAs and PDs. While their output also includes frequency and spatial information their validation focused only on 10 patient cases where only the classification of patterns was compared to the neurophysiology report for accuracy. Here we have expanded the validation work by incorporating more than 300 EEG segments in the evaluation of the algorithms’ outputs. McGraw et al. [22] extended the identification of patterns with the automatic quantification of frequency and spatial extent of LPDs and GPDs while looking at changes in the energy of the time-domain signal with a sliding window. An agreement of above 90% was observed when three clinical experts evaluated the output of the quantification algorithm. While the overall agreement is high, when quantitatively evaluated against the frequency of the events and their spatial extent, the metrics showed a reduced performance. By applying a peak detector on the first derivative instead of the time domain signal, we have improved the results based on the quantitative evaluation. Several other works [23], [24] propose methods for the detection of periodic discharges, but do not include LRDA and GRDA activity and do not evaluate frequency and spatial extent. In this work, we have added to the state-of-the-art by also developing algorithms that target the quantification of LRDA and GRDA patterns.

Our work has several limitations. The evaluation of algorithm performance was based on a limited number of LRDA, GRDA, LPD and GPD segments as we selected only those that had complete consensus on classification between the annotators. While this limits the number of segments, it allows greater accuracy in assessing the frequency and spatial extent. However, we demonstrate similar performance even when using the full dataset with classification disagreement as shown in Table B2 of the supplemental material. The annotation process involved only three human experts from a single medical facility. Since manual annotations were considered the ground truth, a degree of subjectivity is inherent in the evaluation. While we focused on creating adaptive data-driven methods for event identification, all presented algorithms rely to a certain extent on rule-based selection of events which may require additional validation when applied to other datasets. Integrating frequency and spatial extent estimation with deep learning models may enhance algorithm performance and improve the generalizability of results. Fine-tunning pre-trained neural network models with the LRDA, GRDA, LPD, GPD frequency and spatial extent annotations may result in a model with better performance and broader applicability.

## 5. Conclusions

In this study, we proposed several methods for estimating the frequency and spatial extent of rhythmic and periodic epileptiform activity. Our findings indicate that RDA1b-FFT is the most effective for detecting frequency and spatial extent of rhythmic delta activity, while PD2a performs best for periodic discharges. Both methods demonstrated strong agreement with clinical neurophysiologists’ annotations, producing comparable estimates of event frequency and spatial extent.

## Data availability

All data and code are available upon request to the corresponding author.

## List of abbreviations

ACNS: American Clinical Neurophysiology Society
cEEG: Continuous Electroencephalography
CI: Confidence interval
ea-IRR: expert-algorithm inter-rater reliability
ee-IRR: expert-expert inter-rater reliability
EMD: Empirical Mode Decomposition
FFT: Fast Fourier transform
GPD: Generalized periodic discharges
GRDA: Generalized rhythmic delta activity
HHT: Hilbert-Huang Transform
HSA: Hilbert Spectral Analysis
HT: Hilbert Transform
IIC: Intra-class correlation coefficient
IMF: Intrinsic Mode Function
IRR: Inter-rater reliability
LPD: Lateralized periodic discharges
LRDA: Lateralized rhythmic delta activity
MAE: Mean absolute error
RDA: Rhythmic delta activity
RPP: Rhythmic and periodic patterns
PD: Periodic discharges
PA: Pair-wise agreement

