## Supplemental material for "Automated estimation of frequency and spatial extent of periodic and rhythmic epileptiform activity from continuous electroencephalography data"

#### Appendix A – User Interface for Annotators

Figure A1 presents the interface used by the three neurophysiologists to annotate the frequency and spatial extent of LRDA, GRDA, LPD and GPD segments. The frequency of events is reported as the mean value over the 10 seconds of data displayed. The spatial extent is reported as a ratio between 0 and 1. The spatial extent areas are reported as the brain regions defined in the Methods Section that present the event. The annotators went through each event type provided.

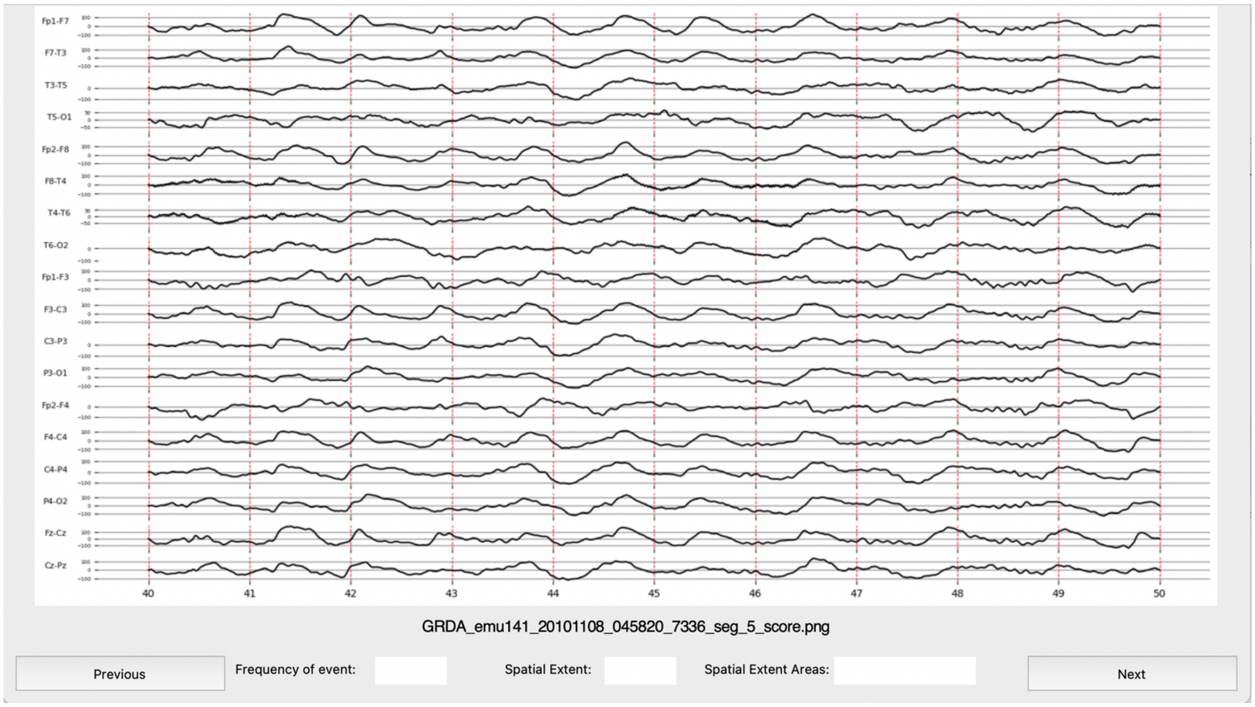

*Figure A1: Interface for the annotation of event frequency and spatial extent of LRDA, GRDA, LPD, GPD segments.*

#### Appendix B – Additional results

*Table B1: Inter-rater reliability results for the intraclass correlation coefficient (ICC) and mean percentage agreement (PA) for expert-expert IRR (ee-IRR) and all expert-algorithms IRR (ea-IRR) for all proposed algorithms along with their confidence intervals (CI) on the EEG segments with 100% agreement on classification of the three experts.*

| | | | ICC | 95% CI | PA | 95% CI |
|---|---|---|---|---|---|---|
| LRDA | | | | | | |
| Frequency of Event | ee-IRR | | 0.88 | [0.80, 0.92] | 0.66 | [0.53,0.77] |
| | ea-IRR | RDA1a-FFT | 0.84 | [0.75, 0.90] | 0.55 | [0.41,0.68] |
| | | RDA1b-FFT | 0.91 | [0.85, 0.94] | 0.60 | [0.47,0.72] |
| | | RDA2-HHT | 0.72 | [0.56, 0.83] | 0.50 | [0.37, 0.63] |
| Spatial Extent | ee-IRR | | 0.80 | [0.68,0.88] | 0.38 | [0.25,0.51] |
| | ea-IRR | RDA1a-FFT | 0.83 | [0.73, 0.89] | 0.31 | [0.20,0.44] |
| | | RDA1b-FFT | 0.85 | [0.77, 0.91] | 0.55 | [0.39,0.64] |
| | | RDA2-HHT | 0.48 | [0.19, 0.68] | 0.39 | [0.27,0.52] |
| GRDA | | | | | | |
| Frequency of Event | ee-IRR | | 0.92 | [0.89,0.94] | 0.74 | [0.66,0.81] |
| | ea-IRR | RDA1a-FFT | 0.95 | [0.94,0.97] | 0.80 | [0.72,0.86] |
| | | RDA1b-FFT | 0.96 | [0.94,0.97] | 0.80 | [0.72,0.86] |
| | | RDA2-HHT | 0.73 | [0.65,0.81] | 0.35 | [0.27,0.44] |
| Spatial Extent | ee-IRR | | 0.60 | [0.46,0.71] | 0.87 | [0.81,0.91] |
| | ea-IRR | RDA1a-FFT | 0.15 | [-0.13,0.27] | 0.43 | [0.35,0.51] |
| | | RDA1b-FFT | 0.66 | [0.55,0.75] | 0.81 | [0.73,0.87] |
| | | RDA2-HHT | 0.51 | [0.34,0.64] | 0.88 | [0.81,0.92] |
| LPD | | | | | | |
| Frequency of Event | ee-IRR | | 0.83 | [0.76,0.88] | 0.63 | [0.53,0.71] |
| | ea-IRR | PD1 | 0.40 | [0.20,0.57] | 0.09 | [0.05,0.16] |
| | | PD2a | 0.80 | [0.73,0.85] | 0.54 | [0.45,0.63] |
| | | PD2b | 0.60 | [0.47,0.71] | 0.14 | [0.09,0.22] |
| Spatial Extent | ee-IRR | | 0.77 | [0.69,0.84] | 0.37 | [0.29,0.45] |
| | ea-IRR | PD1 | 0.65 | [0.53,0.74] | 0.15 | [0.10,0.22] |
| | | PD2a | 0.77 | [0.70,0.84] | 0.43 | [0.34,0.52] |
| | | PD2b | 0.70 | [0.60,0.78] | 0.13 | [0.09,0.19] |
| GPD | | | | | | |
| Frequency of Event | ee-IRR | | 0.86 | [0.81,0.90] | 0.82 | [0.74,0.88] |
| | ea-IRR | PD1 | 0 | [-0.35,0.27] | 0.18 | [0.12,0.13] |
| | | PD2a | 0.61 | [0.47,0.72] | 0.72 | [0.63,0.80] |
| | | PD2b | 0.55 | [0.39,0.67] | 0.03 | [0,0.03] |
| Spatial Extent | ee-IRR | | 0.13 | [-0.19,0.38] | 1 | [0.96,1] |
| | ea-IRR | PD1 | 0.12 | [-0.19,0.36] | 1 | [0.96,1] |
| | | PD2a | 0.13 | [-0.17,0.37] | 0.30 | [0.22,0.39] |
| | | PD2b | 0 | [-0.25,0.33] | 1 | [0.96,1] |

*Table B2: Inter-rater reliability results for the intraclass correlation coefficient (ICC) and mean percentage agreement (PA) for expert-expert IRR (ee-IRR) and all expert-algorithms IRR (ea-IRR) for all proposed algorithms along with their confidence intervals (CI) on all the EEG segments annotated by the raters, regardless of their agreement on segment classification*

| | | | ICC | 95% CI | PA | 95% CI |
|---|---|---|---|---|---|---|
| LRDA | | | | | | |
| Frequency of Event | ee-IRR | | 0.58 | [0.48,0.67] | 0.43 | [0.36,0.49] |
| | ea-IRR | RDA1a-FFT | 0.61 | [0.52,0.7] | 0.27 | [0.21,0.33] |
| | | RDA1b-FFT | 0.69 | [0.62,0.76] | 0.44 | [0.38,0.50] |
| | | RDA2-HHT | 0.50 | [0.38,0.6] | 0.43 | [0.37,0.49] |
| Spatial Extent | ee-IRR | | 0.69 | [0.62,0.76] | 0.22 | [0.17,0.28] |
| | ea-IRR | RDA1a-FFT | 0.75 | [0.69,0.8] | 0.26 | [0.22,0.31] |
| | | RDA1b-FFT | 0.74 | [0.69,0.8] | 0.20 | [0.15,0.26] |
| | | RDA2-HHT | 0.50 | [0.39,0.61] | 0.30 | [0.24,0.35] |
| GRDA | | | | | | |
| Frequency of Event | ee-IRR | | 0.55 | [0.45,0.64] | 0.65 | [0.60,0.71] |
| | ea-IRR | RDA1a-FFT | 0.72 | [0.67,0.77] | 0.24 | [0.20,0.30] |
| | | RDA1b-FFT | 0.72 | [0.67,0.78] | 0.67 | [0.61,0.72] |
| | | RDA2-HHT | 0.46 | [0.35,0.56] | 0.67 | [0.61,0.72] |
| Spatial Extent | ee-IRR | | 0.48 | [0.38,0.58] | 0.53 | [0.48,0.58] |
| | ea-IRR | RDA1a-FFT | 0.57 | [0.49,0.65] | 0.68 | [0.63,0.72] |
| | | RDA1b-FFT | 0.59 | [0.52,0.67] | 0.23 | [0.18,0.28] |
| | | RDA2-HHT | 0.35 | [0.23,0.47] | 0.51 | [0.46,0.57] |
| LPD | | | | | | |
| Frequency of Event | ee-IRR | | 0.69 | [0.62,0.75] | 0.63 | [0.57,0.69] |
| | ea-IRR | PD1 | 0.52 | [0.43,0.61] | 0.14 | [0.10,0.18] |
| | | PD2a | 0.68 | [0.62,0.74] | 0.60 | [0.55,0.66] |
| | | PD2b | 0.59 | [0.51,0.67] | 0.09 | [0.06,0.13] |
| Spatial Extent | ee-IRR | | 0.60 | [0.52,0.68] | 0.43 | [0.38,0.48] |
| | ea-IRR | PD1 | 0.53 | [0.44,0.62] | 0.13 | [0.10,0.16] |
| | | PD2a | 0.68 | [0.62,0.74] | 0.50 | [0.44,0.55] |
| | | PD2b | 0.55 | [0.46,0.63] | 0.12 | [0.09,0.15] |
| GPD | | | | | | |
| Frequency of Event | ee-IRR | | 0.37 | [0.24,0.49] | 0.79 | [0.74,0.84] |
| | ea-IRR | PD1 | -0.04 | [0.25,0.14] | 0.10 | [0.07,0.14] |
| | | PD2a | 0.18 | [0.03,0.33] | 0.77 | [0.71,0.81] |
| | | PD2b | 0.32 | [0.19,0.44] | 0.01 | [0.004,0.03] |
| Spatial Extent | ee-IRR | | 0.28 | [0.13,0.42] | 0.57 | [0.52,0.62] |
| | ea-IRR | PD1 | 0.25 | [0.11,0.38] | 0.76 | [0.72,0.79] |
| | | PD2a | 0.36 | [0.24,0.48] | 0.19 | [0.14,0.23] |
| | | PD2b | 0.25 | [0.11,0.39] | 0.76 | [0.72,0.79] |

*Table B3: MAE (mean absolute error) between the mean human expert annotated value and the algorithms for RDA (rhythmic delta activity) and PD (periodic discharges), along with their 95% CI for all segments included in this work, regardless of the raters' agreement on segment classification.*

| | **Algorithm** | **LRDA** | **GRDA** | **Algorithm** | **LPD** | **GPD** |
|---|---|---|---|---|---|---|
| Frequency of Event [Hz] | RDA1a-FFT | 0.19 [0.13,0.25] | 0.24 [0.19,0.30] | PD1 | 1.19 [1.02,0.36] | 0.98 [0.86,1.11] |
| | RDA1b-FFT | 0.13 [0.09,0.17] | 0.27 [0.21,0.32] | PD2a | 0.42 [0.38,0.45] | 0.15 [0.13,0.18] |
| | RDA2-HHT | 0.13 [0.10,0.16] | 0.46 [0.39,0.52] | PD2b | 0.57 [0.50,0.63] | 1.25 [1.19,1.29] |
| Spatial Extent | RDA1a-FFT | 0.26 [0.22,0.30] | 0.32 [0.28,0.40] | PD1 | 0.53 [0.50,0.56] | 0.01 [0.01,0.01] |
| | RDA1b-FFT | 0.19 [0.16,0.21] | 0.09 [0.07,0.10] | PD2a | 0.17 [0.15,0.19] | 0.40 [0.36,0.44] |
| | RDA2-HHT | 0.14 [0.12,0.17] | 0.07 [0.06,0.09] | PD2b | 0.60 [0.59,0.60] | 0.02 [0.01,0.02] |

### Appendix C – Agreement Matrices

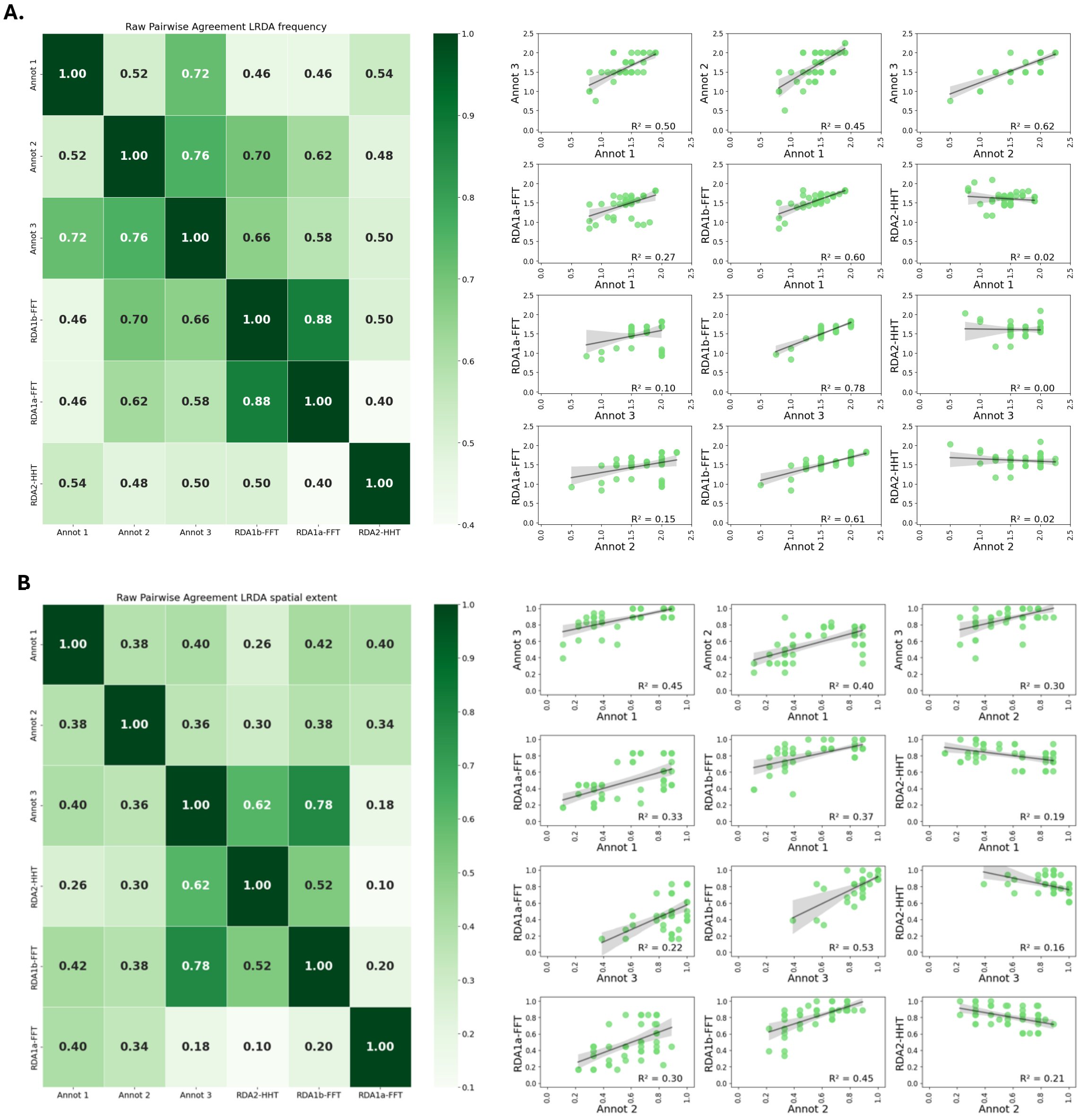


*Figure C1: LRDA raw pair-wise agreement of the annotators and the algorithm along with scatterplots of the annotations. A. Frequency of event. B. Spatial extent.*

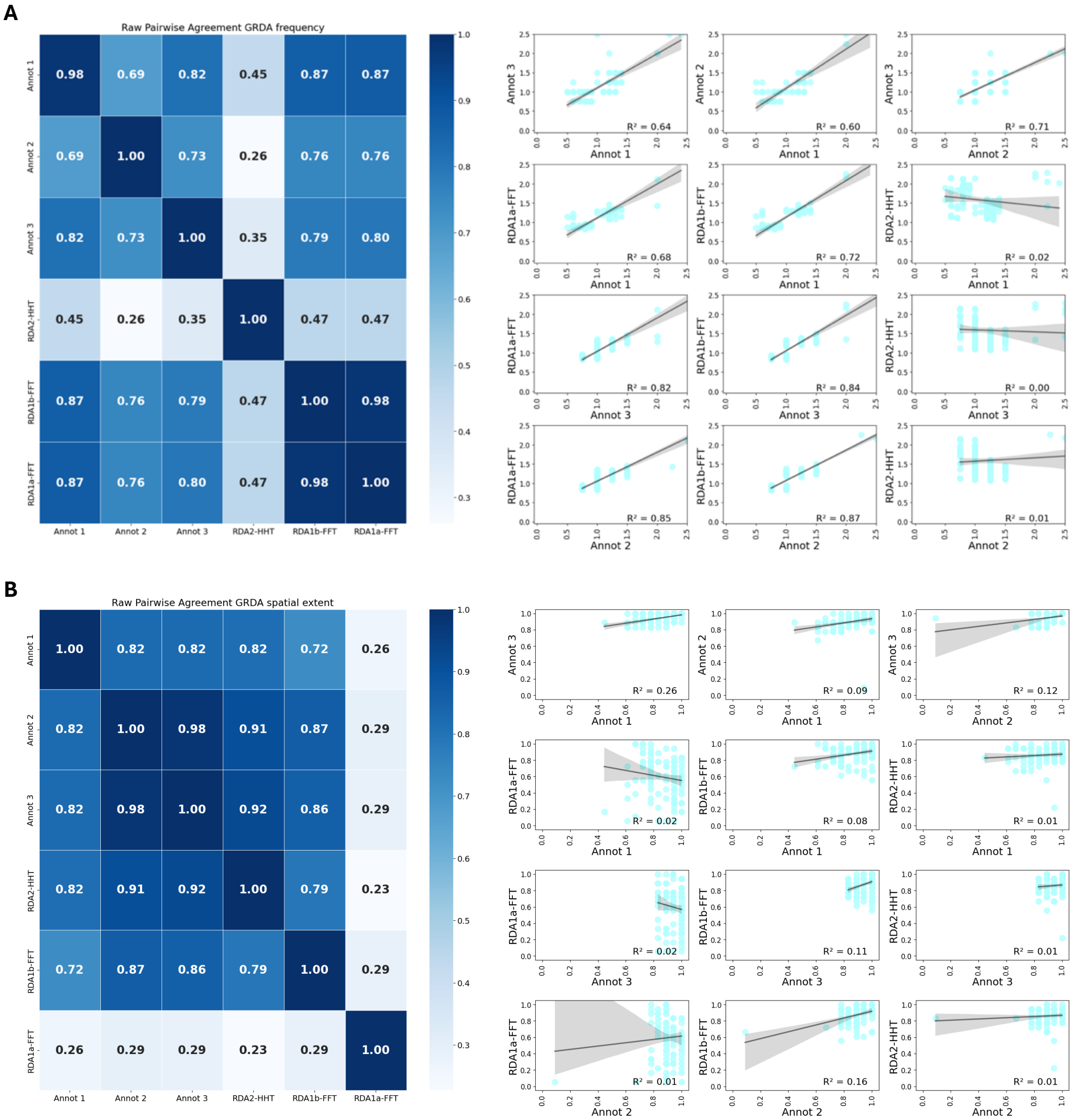

*Figure C2: GRDA raw pair-wise agreement of the annotators and the algorithm along with scatterplots of the annotations. A. Frequency of event. B. Spatial extent.*

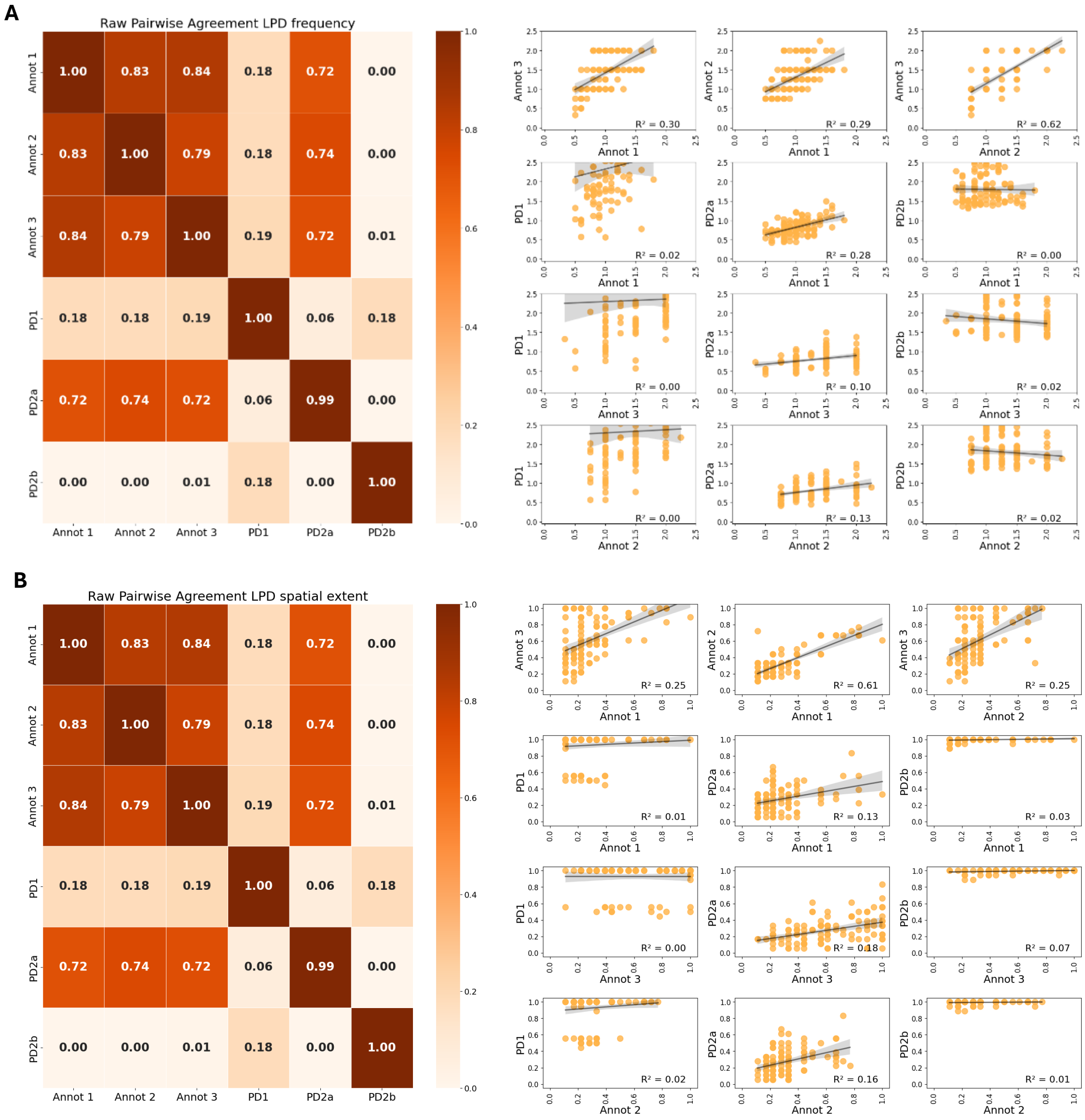


*Figure C3: LPD raw pair-wise agreement of the annotators and the algorithm along with scatterplots of the annotations. A. Frequency of event. B. Spatial extent.*

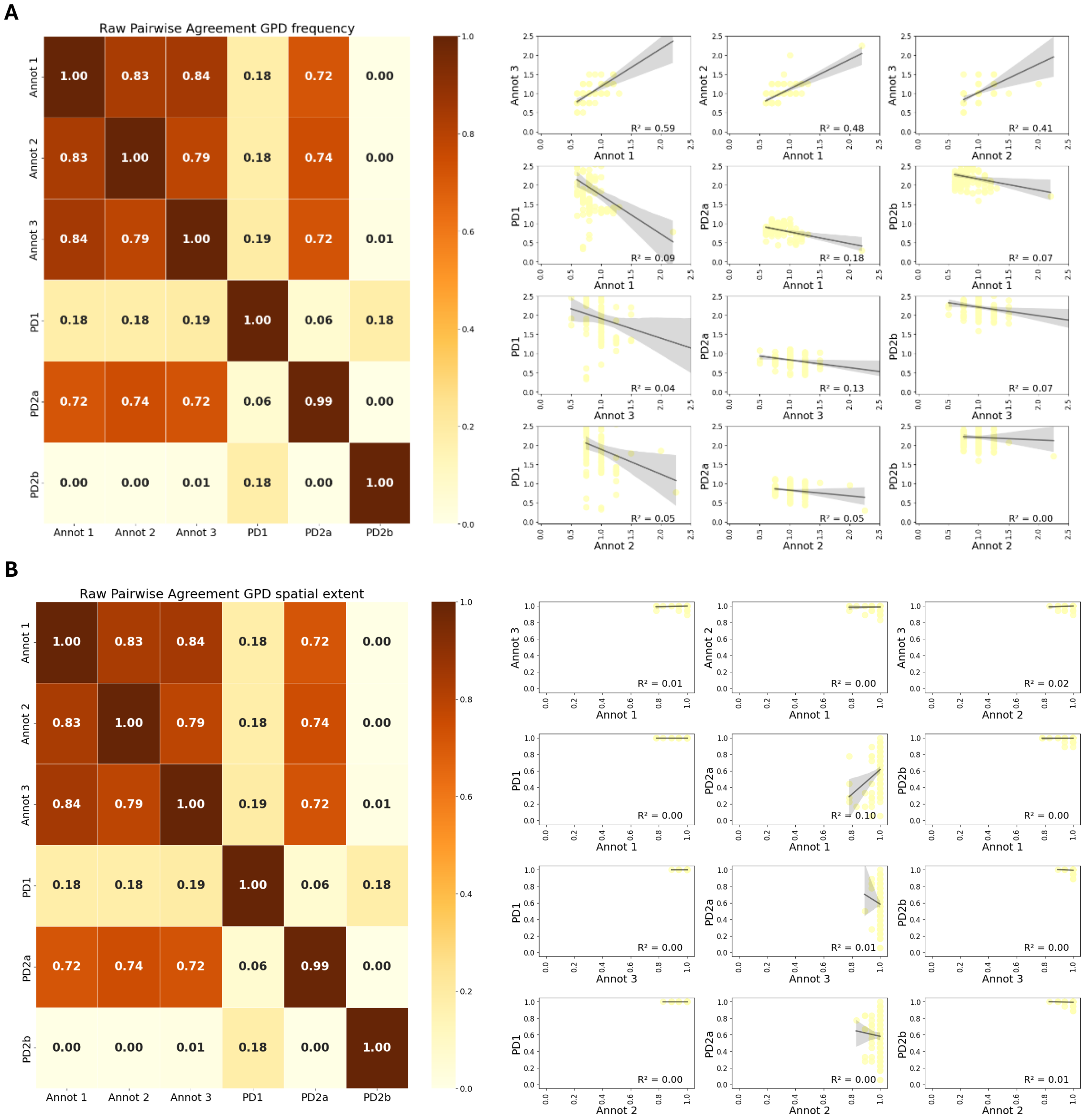


*Figure C4: GPD raw pair-wise agreement of the annotators and the algorithm along with scatterplots of the annotations. A. Frequency of event. B. Spatial extent.*
